# Unmeasured confounding and misclassification in vaccine effectiveness studies using electronic health records (EHRs): an evaluation of a multi country European study (VEBIS-EHR)

**DOI:** 10.1101/2025.04.30.25326722

**Authors:** James Humphreys, Nathalie Nicolay, Toon Braeye, Izaak Van Evercooren, Christian Holm Hansen, Ida Rask Moustsen-Helms, Chiara Sacco, Massimo Fabiani, Jesús Castilla, Iván Martínez-Baz, Ausenda Machado, Patricia Soares, Brechje de Gier, Hinta Meijerink, Susana Monge, Sabrina Bacci, Baltazar Nunes, VEBIS-EHR working group

## Abstract

**Background:** Electronic health record (EHR)-based observational studies can rapidly provide real-world data on vaccine effectiveness (VE), particularly key during the COVID-19 pandemic. However, EHR data may be prone to misclassification and unmeasured confounding, requiring systematic mitigation to ensure robust findings.

**Methods:** In VEBIS-EHR, a retrospective multi-country COVID-19 VE cohort study, we examined unmeasured confounding using a negative control outcome (death not related to COVID-19) and misclassification from varying data extraction intervals. The evaluation spanned two periods (November-December 2023, January-February 2024), encompassing up to 18.7 million individuals across six EU/EEA countries. Vaccine confounding-adjusted hazard ratios (aHRs) were pooled using random-effects meta-analysis.

**Results:** aHRs against non–COVID-19 mortality ranged from 0.35 (95% CI: 0.28–0.44) to 0.70 (0.66–0.73) when comparing vaccinated versus unvaccinated. Delaying EHR data extraction modestly increased the capture of outcome and exposure events, with some variation by vaccination status. Site-level fluctuations in aHRs did not meaningfully alter the overall pooled VE, suggesting stable estimates despite misclassification related to extraction timing.

**Conclusions:** We observed some evidence of unmeasured confounding when using non-COVID-19 deaths as a negative outcome, though the specificity of our negative control must be considered. This result may suggest overestimation of VE, but also the need for further analysis with more specific negative control outcomes and confounding-adjustment techniques. Addressing such confounding using richer data sources and more refined approaches remains critical to ensure accurate, timely VE estimates when using real-world EHR-based data. Extending the delay between the end of observation and data extraction modestly improves the completeness of exposure and outcome data, with limited effect on pooled VE estimates.

**Key Messages:** A Key Messages section should be added after the keywords and before the article’s introduction, with the key messages of the paper made in 3 bullet points that succinctly describe:

- **What your research question was:** Whether vaccine effectiveness estimates based on EHR data are internally valid according to analyses focused on two topics of concern related to EHR-based observational research – unmeasured confounding and misclassification.
- **What you found:** We observed unmeasured confounding in our estimates when using non-COVID19 deaths as negative outcome, and timing of data abstraction from source EHRs was found to have a small influence on the capture and classification of exposure and outcome.
- **Why it is important:** Our results reinforce earlier evidence of the healthy vaccinee phenomenon indicating possible biases in the estimates of vaccine effectiveness estimates obtained from EHR data and that there appears to be little influence of EHR extraction timing on data completeness.

## Introduction

Electronic health records (EHRs) have increasingly been used in epidemiological research to provide extensive real-world data for evaluating health interventions, including in studies estimating vaccine effectiveness (VE) (1). EHRs capture routinely-collected clinical data such as diagnoses, laboratory results, and vaccination events from real-world healthcare settings, gradually proving their utility in large-scale observational epidemiological studies designed to provide timely evidence for public health policy – a factor of particular importance during the COVID-19 pandemic (2–12).

Because EHRs are generally collected for clinical rather than research purposes, studies relying on EHR-derived data for secondary analysis must account for specific sources of potential bias – particularly information bias arising from misclassification of exposures or outcomes (13,14). This makes the original intent of data collection a key consideration; for example, databases designed for billing may be unique in their coding practices, which can render their records less suitable for use in research, surveillance and monitoring. Another potential source of misclassification is incomplete or delayed reporting of exposures (e.g., vaccination events) and outcomes (e.g., cause of death) which, if not monitored and adjusted for, may bias effect estimates drawn from the abstracted data (15–18).

If adjustment for confounders is inadequate, the estimates may be biased whether data are based on EHR or not (19). Investigators relying on the secondary usage of data from EHRs are not generally able to adapt their data scope (e.g., to include additional measures from patients) according to study requirements, which may make accurate effect estimation challenging if data necessary to perform key confounding bias adjustments are not available or linkable (20,21). Studies aiming to evaluate interventions using EHRs, including VE studies, must ensure that the presence and degree of confounding, in particular unmeasured differences in baseline risks between the exposed and unexposed, are sufficiently evaluated and described (22). Considering these and other limitations, the internal validity of estimates based on EHR data has been discussed extensively to date (23,24).

The focus of the present evaluation is an EHR-based VE monitoring platform, the VEBIS project (Vaccine Effectiveness, Burden and Impact Studies) funded by the European Centre for Disease Prevention and Control (ECDC) (25). The VEBIS-EHR study aims to monitor COVID-19 VE in real time using data from population EHR across six EU/EEA countries against severe outcomes of COVID-19 including COVID-19 related hospitalizations and deaths in individuals aged 65+ or other population subgroups (25–30).

In order to assess the validity of VE, estimates produced using data sourced from EHRs we evaluated i) unmeasured confounding in models estimating COVID-19 VE against COVID-19-related deaths using a negative control outcome to assess differences in baseline risks in the exposed versus unexposed, and ii) how the timing of EHR data extraction affects classification of exposure and outcome, and the impact thereof on estimates of COVID-19 VE against COVID-19 related hospitalisation and death.

## Methods

### VEBIS-EHR study

VEBIS-EHR is a retrospective multi-site (currently: Belgium, Denmark, Italy, Norway, Portugal, and Spain (Navarre)) cohort study that aims to monitor autumnal VE against COVID-19-related hospitalisation and death. Further details of the VEBIS-EHR methodology have been described elsewhere (25–30). Briefly, monthly estimates of VE are based on events occurring over rolling eight-week observational periods. Extraction of data from source EHRs is performed at the study site level, typically undertaken 4–6 weeks after the end of each rolling eight-week observation period to allow sufficient time for data consolidation across the respective source databases.

The evaluation spanned two periods (November–December 2023, January–February 2024). The population of interest included individuals eligible for autumn 2023/24 vaccination and resident within each study site, aged 65 years or older and who had completed their primary vaccination series at least 180 days before the start of the 2023/24 vaccination campaign in each respective country, and with no prior COVID-19 vaccination or documented SARS-CoV-2 infection in the 90 days preceding that date, which is defined at the study site level. The exposure was defined as receipt of the autumn 2023 COVID-19 booster vaccine, and time-at-risk as exposed was counted only after 14 days post-vaccination had elapsed. Outcome events of interest were COVID-19 hospitalisation (hospital admission for severe acute respiratory infection with a positive SARS-CoV-2 test from 14 days before to 1 day after admission, or a primary COVID-19 diagnosis) and COVID-19–related death (COVID-19 recorded as cause of death, or unknown cause with death occurring within 30 days of a positive test).

Data were extracted from national or regional electronic health databases by up to six countries which were selected for this evaluation from the seven participating VEBIS-EHR study sites according to data availability and participation at the time of this evaluation (25). Individual-level data from EHRs captured information on the exposure and outcomes of interest, age, sex, region, comorbidities, prior number of booster doses received by autumn 2023 vaccine campaign start and, in some study sites, nationality and socioeconomic status, which were used to adjust the estimates for confounding. Adjusted hazard ratios (aHR) with 95% confidence intervals (CI) were estimated using Cox proportional hazards models with time-varying exposure according to time since vaccination (14-89, 90-179, and 180+ days since vaccination) and using calendar time as the underlying time scale. Analyses were stratified by age group (65-79 years-old versus ≥80 years-old). A two-stage meta-analysis was conducted, with each study site calculating site-specific aHRs and CIs and, where required, site level estimates were pooled via a random-effects meta-analysis (Paule-Mantel method) to yield study-level aHR estimates with heterogeneity calculated for each pooled estimate to reflect the proportion of variance attributable to heterogeneity as opposed to chance, represented as I^2^ (31).

### Unmeasured confounding using a negative control outcome

To detect unmeasured confounding in estimates of VE against COVID-19-related death produced according to the above-described methods, we applied the Cox regression model without pooling to a negative control outcome defined as death *not* related to COVID-19 intended to represent the inverse of the main mortality definition (COVID-19 not reported in cause of death and no record of a positive test indicating SARS-CoV-2 infection within the 30 days preceding death). The study period between 1 November to 25 December 2023, after the start of the autumn 2023 vaccination campaign, was selected to include the maximum number of study sites according to their data access, and included four VEBIS-EHR study sites which had access to death databases and could adapt death definitions at the time of the evaluation (Denmark, Navarra, Norway, and Portugal). Where estimates of aHR against non-COVID-19-related death meaningfully differ from the expected threshold of no difference in risk (aHR = 1), including upper 95% confidence intervals that do not cross this threshold, they should be interpreted as an indication of unmeasured confounding, as we do not expect COVID-19 vaccination status to influence the risk of death unrelated to COVID-19, assuming no substantial degree of misclassification in the negative control outcomes (32).

### Underreporting of outcomes and misclassification of vaccine status due to delayed recording

In a separate analysis designed to evaluate the impact of extraction delay on data consolidation, we described the number of eligible participants and relevant outcome events (hospitalisations or deaths related to COVID-19) by vaccination status and study site using three different consolidation periods: 4– 6 weeks post-observation period (the ‘usual’ delay (25)), 8–10 weeks post-observation period (‘intermediate’ delay), and 26–32 weeks post-observation period (‘longest’ delay) (Figure 1). We calculated aHR for each extraction delay and age group according to the VEBIS-EHR methods described above, to assess the impact of these delays on estimates both at the site and pooled level (25). The study period selected was 1 January to 25 February 2024, as it permitted the re-extraction and analysis of data up to 32 weeks after study end during the course of the evaluation, and according to data access constraints which would prevent the participation of Norway in later observation periods. All six of the VEBIS-EHR study sites (Belgium, Denmark, Navarra, Norway, Portugal, and Italy) who participated in the wider evaluation provided data for this analysis.

**FIGURE 1.**
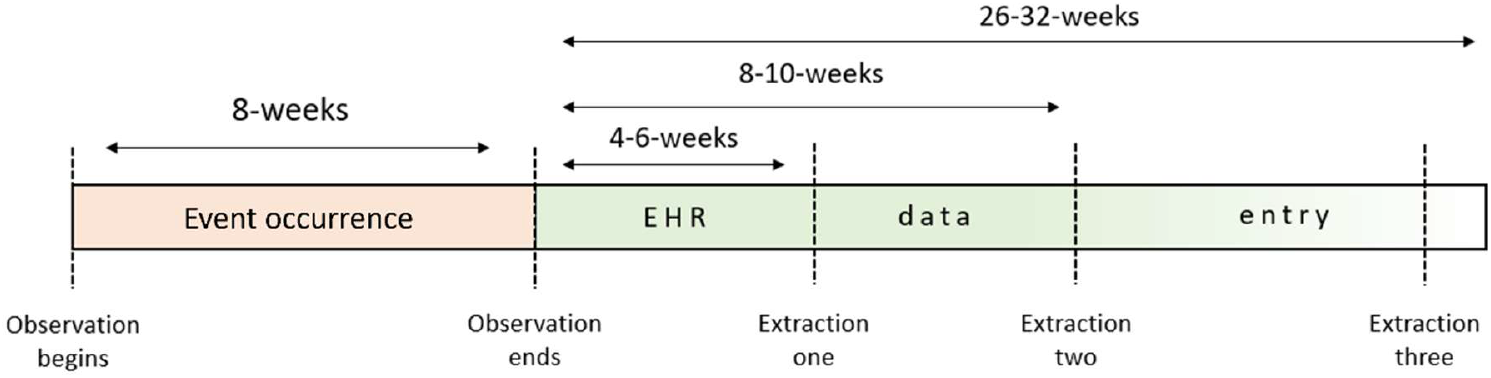
DIAGRAM OF DATA EXTRACTION SCHEMA ILLUSTRATING USUAL (4-6 WEEK), INTERMEDIATE (8-10 WEEK) AND LATE (26-32) EXTRACTIONS VERSUS HEALTHCARE EVENT OCCURRENCE AND PROBABLE ELECTRONIC HEALTH RECORD DATA ENTRY PERIOD.

## Results

### Unmeasured confounding using a negative control outcome

Among the four study sites participating in this exercise, there were up to 4.3 million eligible individuals abstracted from source EHRs who contributed 7.7 million person-months at risk throughout the study period. A total of 8,811 relevant death events were identified, of which 291 were classified as related versus 8,520 unrelated to COVID-19 (Figure 2). The majority (71%) of all events occurred among those aged 80 years plus.

**FIGURE 2.**
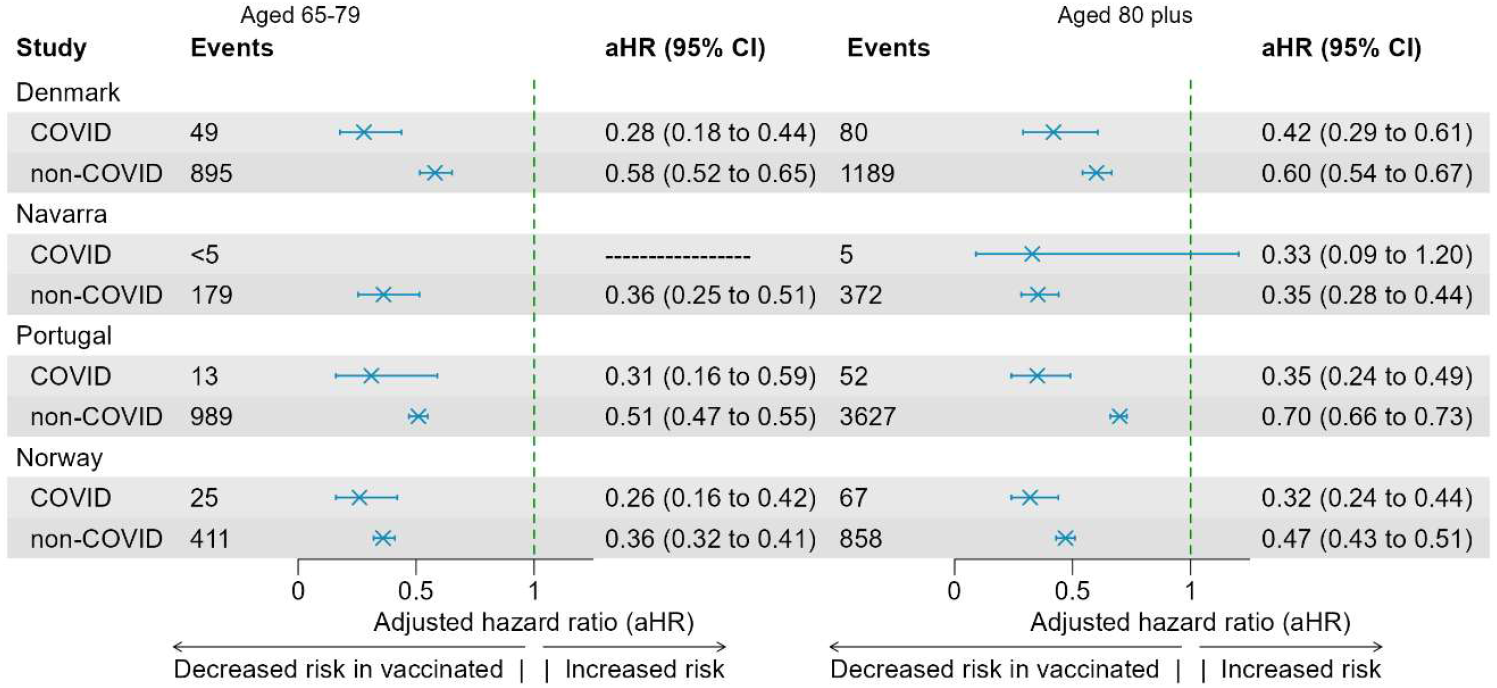
ESTIMATES OF CONFOUNDER-ADJUSTED HAZARD RATIOS AGAINST COVID-19-RELATED DEATH (COVID) AND THE NEGATIVE OUTCOME CONTROL (NON-COVID) AMONG THOSE AGED 65-79 (LEFT) AND 80 PLUS (RIGHT).

The adjusted hazard ratio (aHR) of vaccination for non–COVID-19 mortality was consistently lower than the null effect threshold (aHR=1.0) in all study sites, indicating that vaccinated individuals had a reduced risk of death unrelated to COVID-19 compared to the unvaccinated population. Among those aged 65– 79 years, aHR ranged from 0.36 (95% CI: 0.25–0.51) to 0.58 (95% CI: 0.52–0.65), and from 0.35 (95% CI:

0.28–0.44) to 0.70 (95% CI: 0.66–0.73) among those aged ≥80 years-old (Figure 2). Confidence intervals (95% CI) for estimates of aHR of vaccination against non-COVID-19-related death did not cross the highlighted threshold (aHR) in any study site or age group.

### Underreporting of outcomes and misclassification of vaccine status due to delayed recording

Extracting data 4-6 weeks after the end of the observation period (25^th^ February 2025) yielded a baseline total of 18.7 million eligible individuals, among whom 4,389 relevant COVID-19-related hospitalisations and 793 COVID-19-related deaths were identified and formed the reference for comparison with later data extractions (Annex, Tables 1 to 3).

**TABLE 1.**
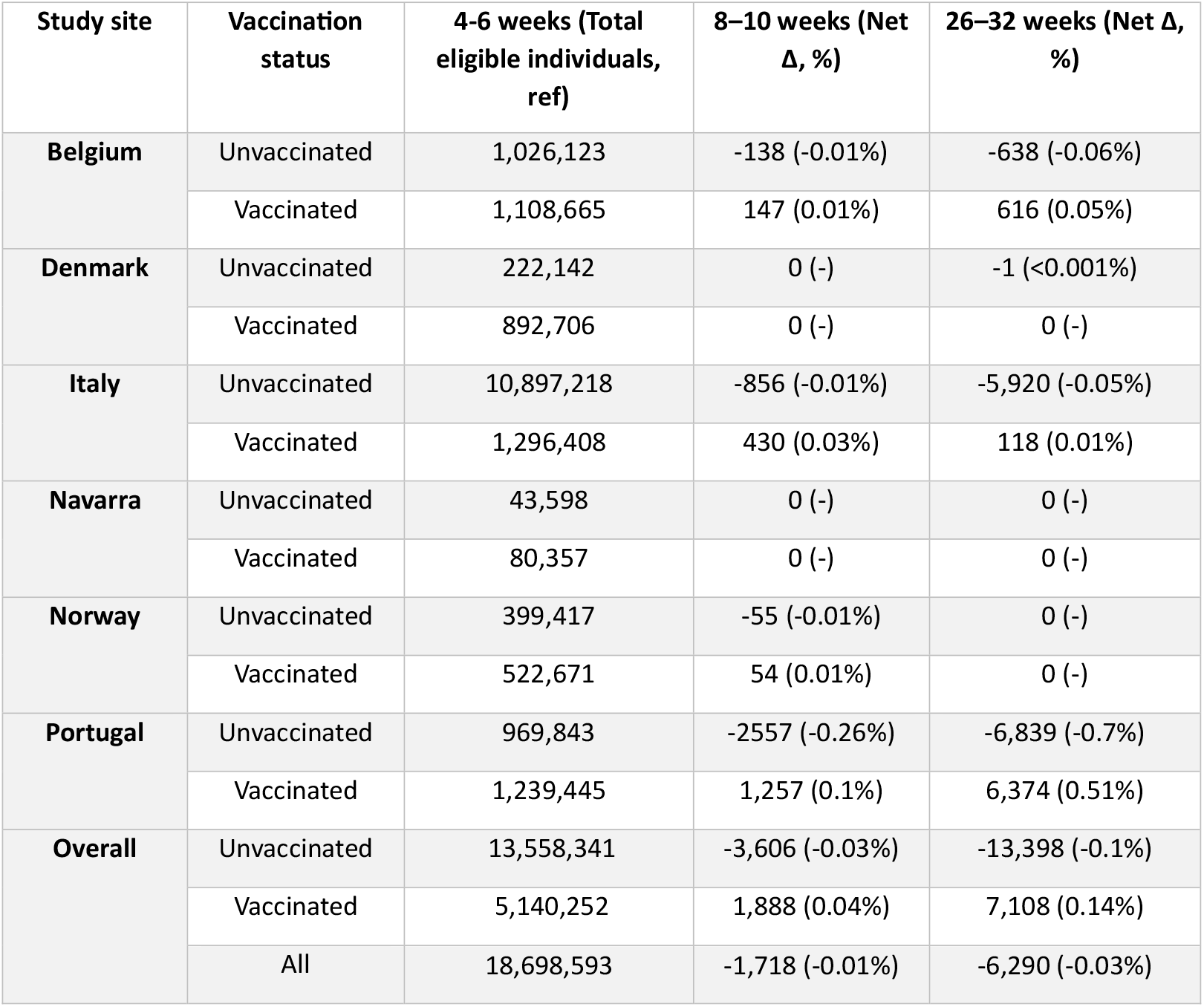
TOTAL INITIAL AND NET PROPORTIONAL CHANGE IN RECORD COUNT STRATIFIED BY VACCINATION STATUS AND STUDY SITE CALCULATED USING EACH SUBSEQUENT DATA EXTRACTION VERSUS USUAL DELAY (4-6 WEEKS AFTER STUDY END ON 25^TH^ FEBRUARY, BETWEEN 24^TH^ MARCH AND 7^TH^ APRIL 2024).

| Study site | Vaccination status | 4-6 weeks (Total eligible individuals, ref) | 8-10 weeks (Net Δ, %) | 26-32 weeks (Net Δ, %) |
| --- | --- | --- | --- | --- |
| Belgium | Unvaccinated | 1,026,123 | -138 (-0.01%) | -638 (-0.06%) |
|  | Vaccinated | 1,108,665 | 147 (0.01%) | 616 (0.05%) |
| Denmark | Unvaccinated | 222,142 | 0 (-) | -1 (<0.001%) |
|  | Vaccinated | 892,706 | 0 (-) | 0 (-) |
| Italy | Unvaccinated | 10,897,218 | -856 (-0.01%) | -5,920 (-0.05%) |
|  | Vaccinated | 1,296,408 | 430 (0.03%) | 118 (0.01%) |
| Navarra | Unvaccinated | 43,598 | 0 (-) | 0 (-) |
|  | Vaccinated | 80,357 | 0 (-) | 0 (-) |
| Norway | Unvaccinated | 399,417 | -55 (-0.01%) | 0 (-) |
|  | Vaccinated | 522,671 | 54 (0.01%) | 0 (-) |
| Portugal | Unvaccinated | 969,843 | -2557 (-0.26%) | -6,839 (-0.7%) |
|  | Vaccinated | 1,239,445 | 1,257 (0.1%) | 6,374 (0.51%) |
| Overall | Unvaccinated | 13,558,341 | -3,606 (-0.03%) | -13,398 (-0.1%) |
|  | Vaccinated | 5,140,252 | 1,888 (0.04%) | 7,108 (0.14%) |
|  | All | 18,698,593 | -1,718 (-0.01%) | -6,290 (-0.03%) |

The total number of participants varied slightly with increasing delays to extraction. Decreases of 1,718 eligible participants (−0.01%), and 6,290 (−0.03%) were observed, respectively, between the intermediate and latest delay extractions versus the reference extract (Annex, Table 1). When stratified by vaccination status there was some evidence of a differential impact, as numbers of vaccinated individuals increased while unvaccinated counts declined.

Compared with the reference extraction and pooled across all study sites, there were an additional 144 (3.28%) and 148 (3.37%) COVID-19–related hospitalisations, and an additional 14 (1.77%) and 15 (1.89%) COVID-19 related deaths, respectively, in the intermediate and the latest extracts (Annex, Tables 2 and 3). When stratified by vaccination status, delayed extractions generally led to greater increases in the number of hospitalisations among vaccinated individuals (up to +8.6%) compared with unvaccinated individuals (up to +2.1%). Fluctuations in death counts had an opposing pattern, with a greater degree of increase among those who were unvaccinated. Most additional hospitalisations were observed in Portugal, where reported events increased by over 100% among vaccinated individuals and over 130% in the unvaccinated cohort in later extracts (Annex, Table 2). In contrast, increased COVID-19–related deaths were largely accounted for by Italy, which showed incremental increases of 2.69% to 4.18% in events recorded at the later extraction intervals (Annex, Table 3).

**TABLE 2.**
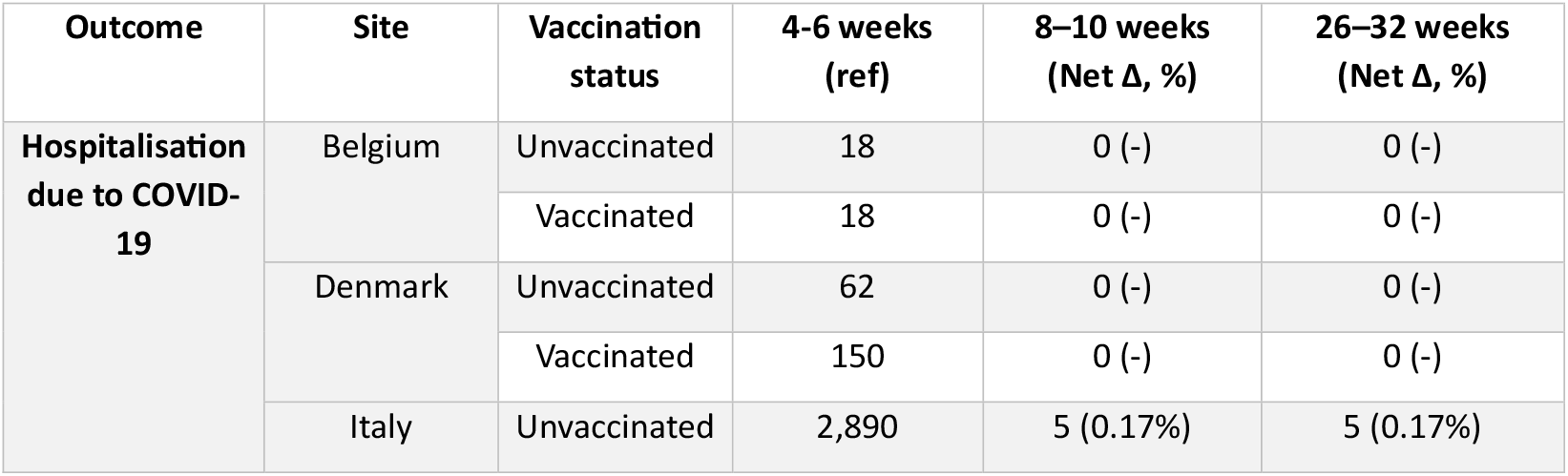
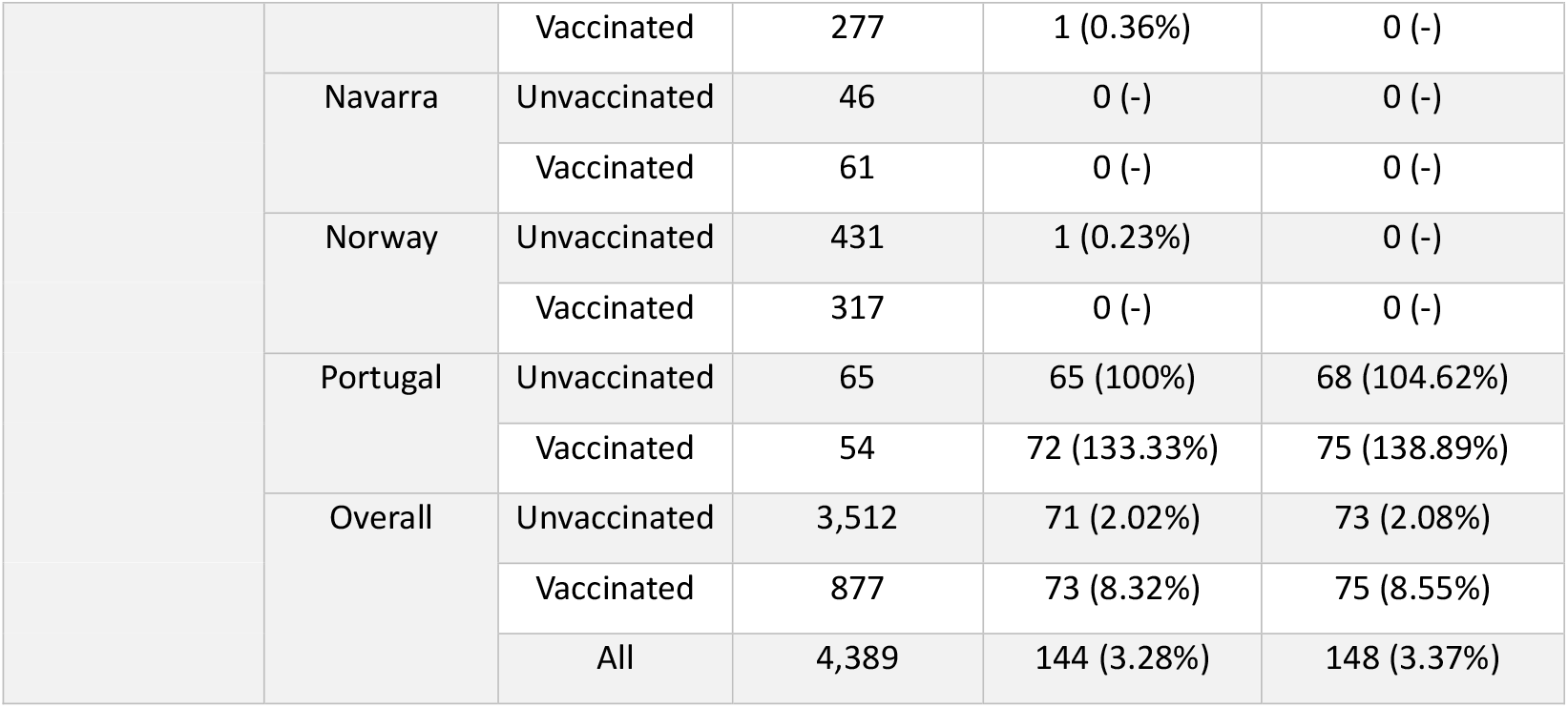
TOTAL INITIAL AND NET PROPORTIONAL CHANGE IN EVENT (HOSPITALISATIONS RELATED TO COVID-19) COUNT STRATIFIED BY VACCINATION STATUS AND STUDY SITE, CALCULATED USING EACH SUBSEQUENT DATA EXTRACTION VERSUS USUAL DELAY (4-6 WEEKS AFTER STUDY END ON 25^TH^ FEBRUARY, BETWEEN 24^TH^ MARCH AND 7^TH^ APRIL 2024).

**TABLE 3.** TOTAL INITIAL AND NET PROPORTIONAL CHANGE IN EVENT (DEATH RELATED TO COVID-19) COUNT STRATIFIED BY VACCINATION STATUS AND STUDY SITE, CALCULATED USING EACH SUBSEQUENT DATA EXTRACTION VERSUS USUAL DELAY (4-6 WEEKS AFTER STUDY END ON 25^TH^ FEBRUARY, BETWEEN 24^TH^ MARCH AND 7^TH^ APRIL 2024).

| Outcome | Site | Vaccination status | Total (ref) | 8–10 weeks (Net Δ, %) | 26–32 weeks (Net Δ, %) |
| --- | --- | --- | --- | --- | --- |
| Death related to COVID-19 | Belgium | Unvaccinated | Not reported |  |  |
|  |  | Vaccinated | Not reported |  |  |
|  | Denmark | Unvaccinated | 35 | 0 (-) | 0 (-) |
|  |  | Vaccinated | 57 | 0 (-) | 0 (-) |
|  | Italy | Unvaccinated | 335 | 9 (2.69%) | 14 (4.18%) |
|  |  | Vaccinated | 30 | 0 (-) | 0 (-) |
|  | Navarra | Unvaccinated | 5 | 0 (-) | 0 (-) |
|  |  | Vaccinated | 9 | 0 (-) | 0 (-) |
|  | Norway | Unvaccinated | 64 | 1 (1.56%) | 0 (-) |
|  |  | Vaccinated | 49 | 0 (-) | 0 (-) |
|  | Portugal | Unvaccinated | 111 | 2 (1.8%) | -1 (-0.9%) |
|  |  | Vaccinated | 97 | 2 (2.06%) | 2 (2.06%) |
|  | Overall | Unvaccinated | 551 | 12 (2.18%) | 13 (2.36%) |
|  |  | Vaccinated | 242 | 2 (0.83%) | 2 (0.83%) |
|  |  | All | 792 | 14 (1.77%) | 15 (1.89%) |

Among individuals aged 65–79 years, the estimated aHR of vaccination against COVID-19–related hospitalisation remained stable across the usual (4–6 weeks), intermediate (8–10 weeks), and latest (26–32 weeks) data extracts for most study sites (Figure 3). An exception was observed in Portugal, where vaccine aHR decreased from 0.56 (95% CI: 0.27–1.17) to 0.4 (95% CI: 0.23–0.7) between the usual and intermediate delays, then remained near this level based on the latest extract. After pooling estimates from all study sites using a random-effects model, pooled vaccine aHR slightly decreased from 0.53 (95% CI: 0.45–0.61) under the usual extraction to 0.52 (95% CI: 0.45–0.6) based on the intermediate extract, and further slightly still to 0.51 (95% CI: 0.44–0.59) based on the latest extract.

**FIGURE 3.**
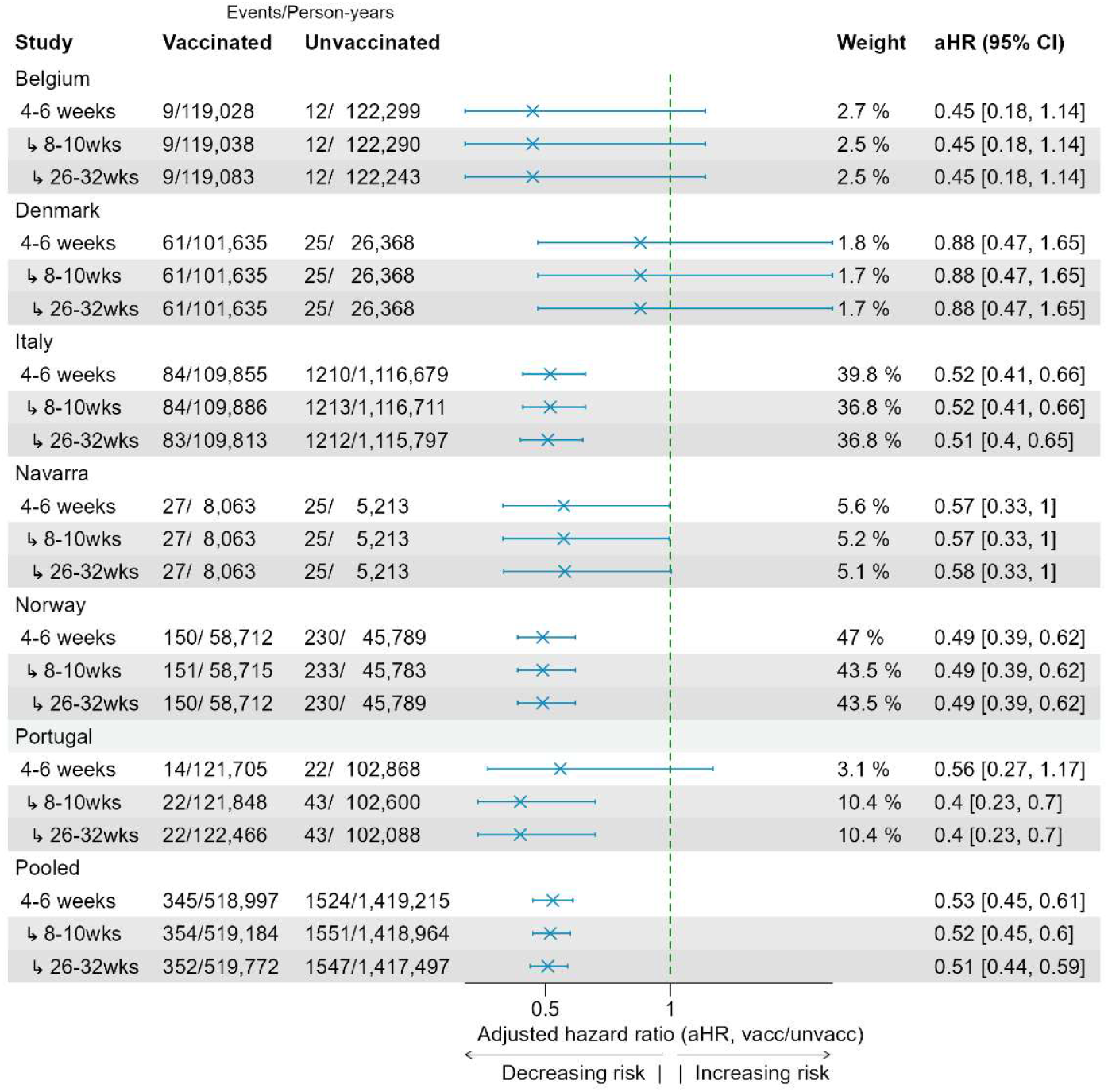
ESTIMATES OF CONFOUNDER-ADJUSTED HAZARD RATIOS AGAINST HOSPITALISATION DUE TO COVID-19 STRATIFIED BY DELAY GROUP AMONG THOSE AGED 65-79.

Among those ≥80-years, vaccine aHR against hospitalisation was also generally consistent across data extracts in most sites (Figure 4). In Portugal, however, aHR fluctuated between 0.49 (95% CI: 0.3–0.78) based on the usual delay to 0.77 (95% CI: 0.56–1.06) in the intermediate delay, remaining stable thereafter based on the latest extract. When pooling estimates from all study sites, the overall vaccine aHR estimate increased slightly from 0.63 (95% CI: 0.55–0.71) to 0.65 (95% CI: 0.58–0.73) in the intermediate extract, remaining stable thereafter.

**FIGURE 4.**
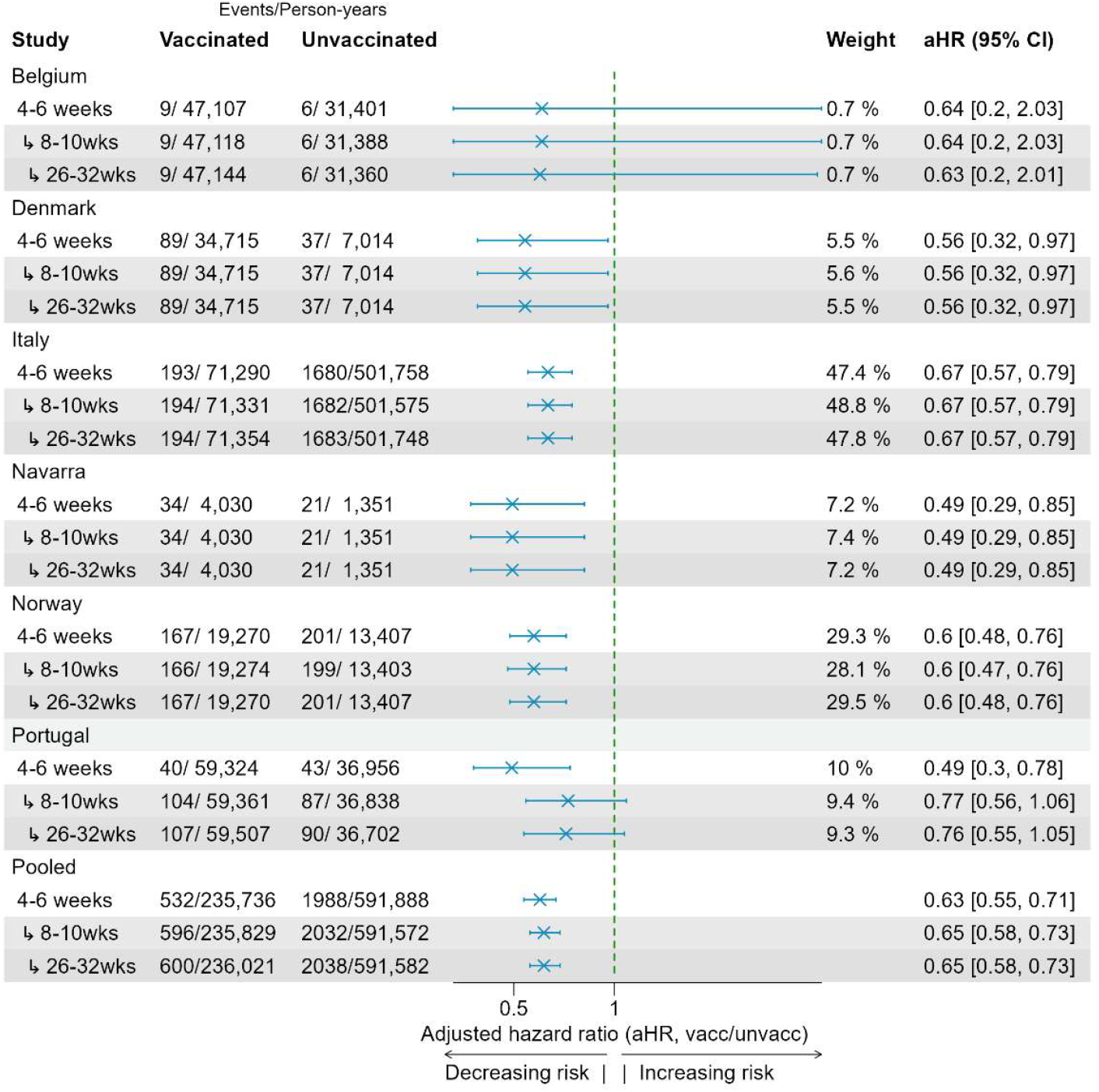
ESTIMATES OF CONFOUNDER-ADJUSTED HAZARD RATIOS AGAINST HOSPITALISATION DUE TO COVID-19 STRATIFIED BY DELAY GROUP AMONG THOSE AGED 80 PLUS.

In the models estimating vaccine aHR against COVID-19–related death in participants aged 65–79 years, fluctuations were no greater than 0.02 in terms of absolute aHR, though estimates were often not at all changed, when compared across the three extracts. As a result, the pooled estimate for this age group and outcome remained essentially unchanged (Figure 5).

**FIGURE 5.**
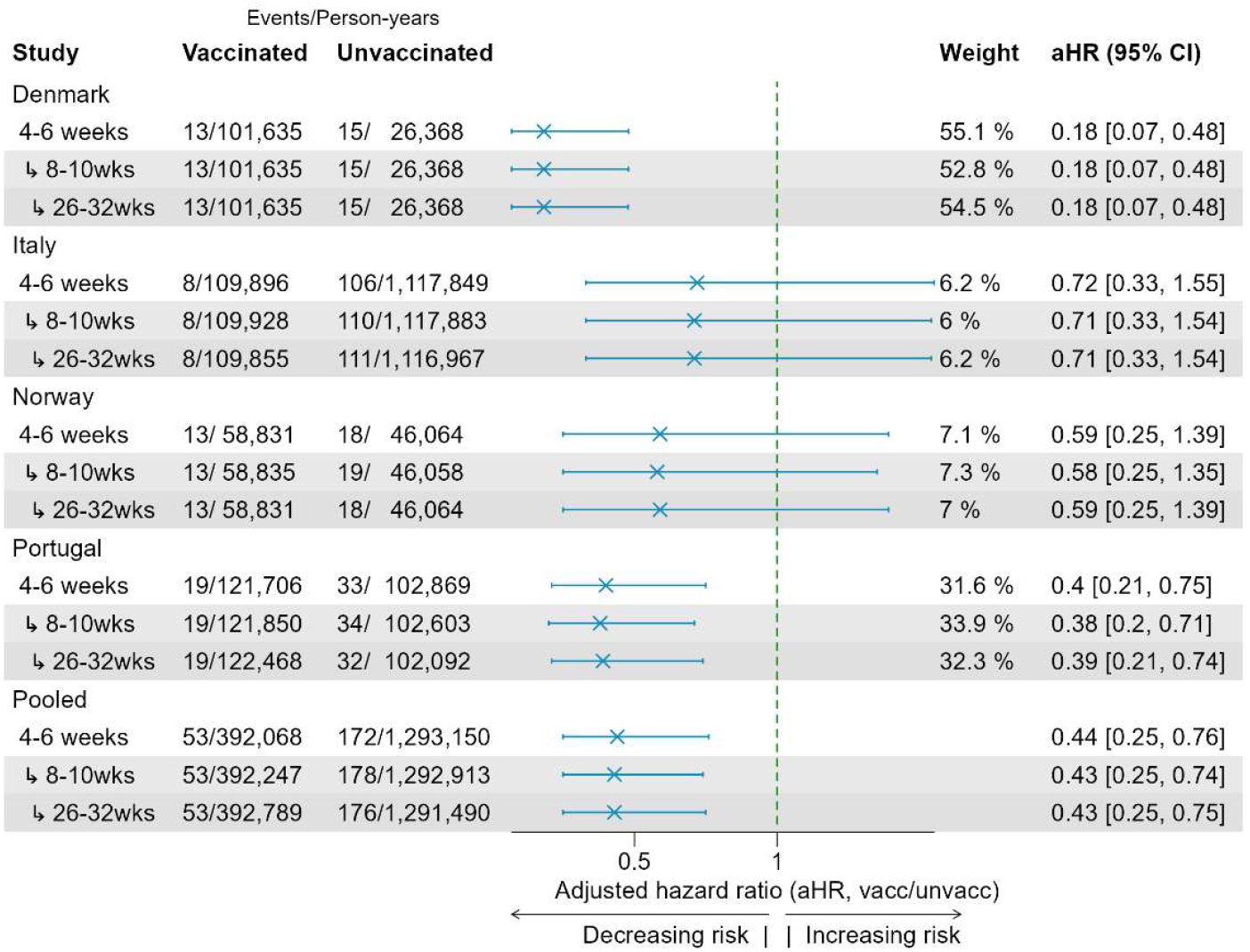
ESTIMATES OF CONFOUNDER-ADJUSTED HAZARD RATIOS AGAINST DEATH RELATED TO COVID-19 STRATIFIED BY DELAY GROUP AMONG THOSE AGED 65-79.

For individuals aged 80 years and older, some study sites reported shifts in vaccine aHR for COVID-19– related death using longer extraction delays (Figure 6). In Italy, estimates of vaccine aHR showed a steady decline from 0.60 (95% CI: 0.37–0.95) using data extracted with the usual delay (4-6 weeks), to 0.58 (95% CI: 0.36–0.92) based on an intermediate delay (8-10 weeks), and finally to 0.55 (95% CI: 0.35– 0.88) based on the data extracted after the longest delay (26-32 weeks). In contrast, Navarra reported that vaccine aHRs that remained consistent between the usual and intermediate extracts at 0.53 (95% CI: 0.16–1.77), but which rose slightly in the latest extraction to 0.58 (95% CI: 0.19–1.77). Because Italy accounted for 27–30% of the overall pooled estimate weighting, reflecting its large number of events and person-months, the gradual decrease seen in Italy was similarly evident at the pooled level. The pooled aHR estimates for death in this older age group remained steady in subsequent extractions.

**FIGURE 6.**
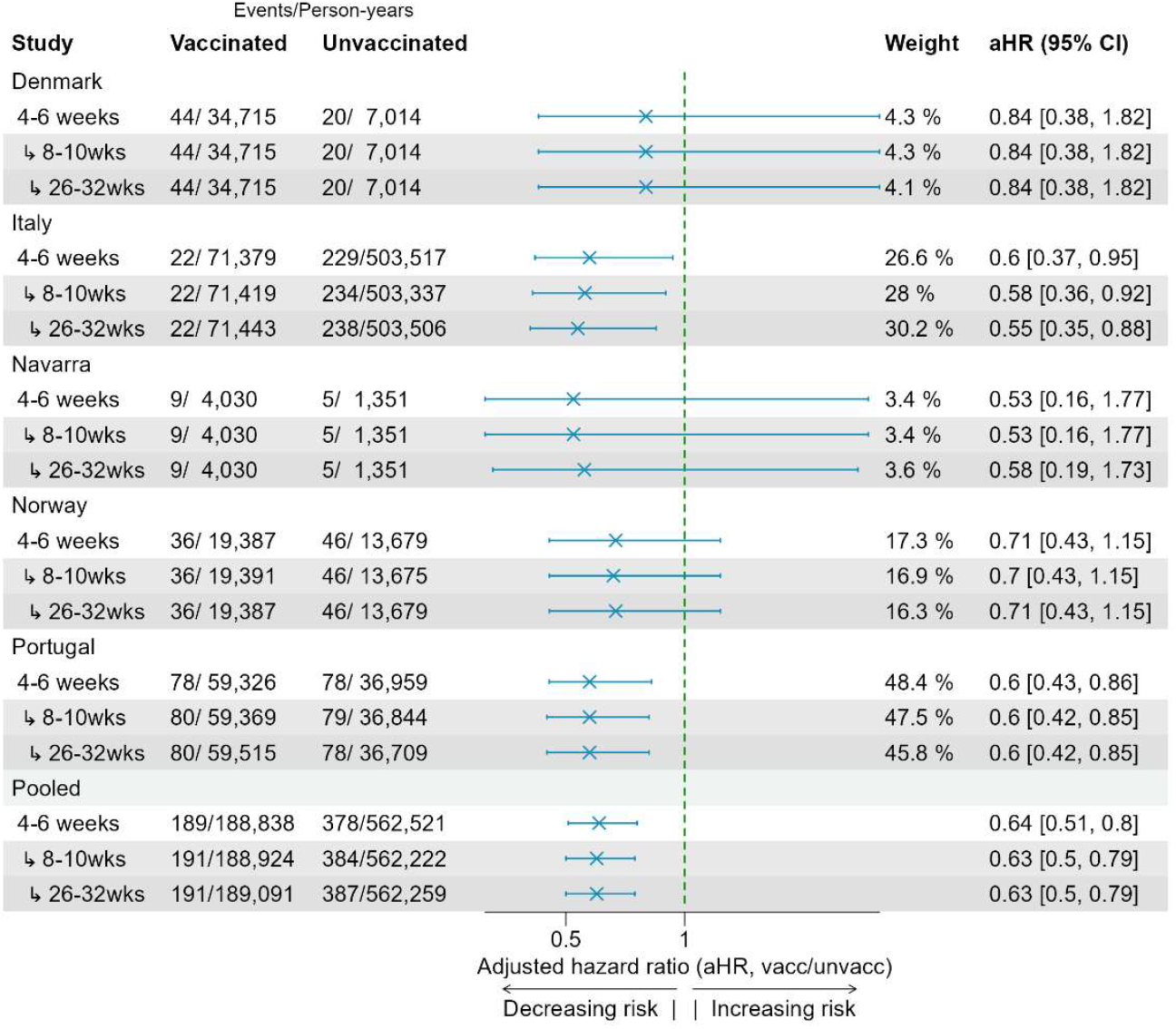
ESTIMATES OF CONFOUNDER-ADJUSTED HAZARD RATIOS AGAINST DEATH RELATED TO COVID-19 STRATIFIED BY DELAY GROUP AMONG THOSE AGED 80 PLUS.

There were no substantial changes in heterogeneity (I^2^) between study sites in any of the four models across both outcomes (hospitalisation or death related to COVID-19) and age groups (65-79 or 80+). Where estimates at the study site level fluctuated, there was generally an increase in the precision of estimates based on data extracted later, likely due to a greater number of events in these extracts versus earlier delays.

## Discussion

Using a negative control outcome, we found that unmeasured confounding may influence VEBIS-EHR’s vaccine effectiveness (VE) estimates. Although data extraction timing can affect the completeness of records, especially for hospitalisations, though these effects appear limited to certain study sites and do not materially alter the pooled VE.

Our negative control outcome analysis demonstrated that adjusted hazard ratios for non-COVID-19 mortality consistently fell below the null, indicating a 30-65% reduced risk of non-COVID-19 deaths among vaccinated individuals. This finding suggests a “healthy-vaccinee effect” and/or the presence of extremely frail individuals in unvaccinated cohorts (33-39). Such patterns, previously noted in influenza studies, imply that individuals who are severely frail or in end-of-life care often do not receive vaccinations and may die earlier, thereby artificially elevating VE estimates if not properly adjusted (34-37). Approaches to mitigating this bias include measuring frailty or health-seeking behaviours more precisely, restricting cohorts to individuals with better prognoses, and accounting for baseline mortality risk (34-36). Although VEBIS-EHR excludes long-term care residents and adjusts for comorbidities, our results suggest these strategies may not fully capture residual frailty. Future evaluations will therefore examine end-of-life status and health-seeking indicators to address confounding further.

Another potential explanation is the choice of negative control outcome. Although we defined non-COVID-19 deaths as having no mention of COVID-19 in the cause of death and no recent positive test, this may not be sufficiently specific (e.g., there could be unrecognised post-COVID-19 complications). Nonetheless, other investigators who used highly specific outcomes (e.g., bone fractures) have still seen spurious protection estimates (39-41). This strengthens the case that unmeasured confounding is not merely an artifact of choosing a less specific negative control outcome.

Addressing unmeasured confounding will likely require richer data sources, more thorough confounder evaluation, and additional analytic methods. Greater data sharing—for instance, through the European Health Data Space—could facilitate linking healthcare records with other sectors, thereby capturing more detailed information on health and behaviours (42,43). Novel data-engineering approaches, including machine learning tools to extract relevant details from unstructured text, may also help identify and exclude individuals who are ineligible or exceptionally frail, thus improving the validity of VE estimates.

In examining data extraction timing, we observed that extending the delay between observation and extraction modestly increased the number of recorded outcomes in some study sites, especially Portugal. This suggests that records initially classified as unvaccinated or without hospitalisations may later be updated as vaccinated or hospitalised. In Italy and Portugal, unvaccinated cohorts shrank in tandem with increases in vaccinated cohorts, pointing to delayed vaccination data entry, although this pattern was not uniform across all sites. Site-specific variability in data completion implies that local validation and sensitivity analyses are critical when interpreting EHR-based VE studies.

Notably, despite these site-level differences, pooled VE estimates remained stable. This finding supports the current 4-6-week data extraction window as a reasonable compromise between timeliness and completeness. Shorter windows (e.g., under four weeks) would likely introduce more misclassification, though we did not formally investigate that scenario. Our results have prompted ongoing analyses within VEBIS-EHR to determine whether identifiable patterns of delayed reporting can guide adjustments to VE estimates.

### Strengths and limitations

A major strength of our study is its multi-country design, which uses a common protocol and a large, representative dataset of routine healthcare interactions from multiple European nations—enhancing both comparability and external validity. However, differences in data entry or coding practices across sites may limit uniformity, as suggested by our observations of delayed event reporting. Additionally, while negative control outcomes can signal unmeasured confounding, they cannot specify its exact sources. Non-COVID-19 mortality may also be insufficiently specific to serve as an ideal control. Future analyses should test more-specific negative outcomes and consider negative exposures, or double negative control (44) as complementary methods.

### Implications for COVID-19 vaccine monitoring and future research

Near real-time COVID-19 VE monitoring is increasingly valuable for public health decision-making, yet few multi-country EHR-based systems have been evaluated, and none, to our knowledge, have investigated how data extraction timing might affect VE. Our results underscore the importance of addressing unmeasured confounding by further probing underlying population differences. This may involve identifying more-appropriate negative control outcomes, employing techniques like negative control exposures or instrumental variables (45-48), and expanding data linkage to capture additional determinants (e.g., health-seeking behaviours, socio-economic factors). Alternative study designs such as self-controlled case series, which can reduce biases arising from inter-individual differences in baseline health, also warrant exploration (49).

As EHR-based monitoring systems mature, more granular data might become available, allowing enhanced confounder control and, by extension, more robust VE estimates. In the interim, the relative stability of pooled estimates despite delayed reporting is reassuring. A 4-6-week extraction window appears adequate, although site-specific patterns of data entry should be closely tracked and longer delays considered as part of sensitivity analyses—particularly in settings with known reporting lags.

## Conclusions

These findings demonstrate that, in VEBIS-EHR, data extraction timing has a modest influence on classification of exposures and events but does not compromise pooled VE estimates. However, negative control outcome analysis revealed notable unmeasured confounding that likely inflates VE results; although the specificity of the chosen negative control outcome remains a factor. Future work should address this confounding through improved data collection, linkage, and analytic strategies, while remaining alert to site-specific differences in data capture.

## Data Availability

Authors cannot share the data used for this study, access to which should be requested from the data owner institutions following their respective procedures.

## Acknowledgments

The authors would like to thank all of those participating in data collection and the production of estimates of VE as without their work these results wouldn’t be available to the scientific community and public.

## Funding

All public health organizations involved received funding from the European Centre for Disease Prevention and Control (ECDC) implementing Framework Contract [ECDC/2021/018] ‘Vaccine effectiveness and impact of COVID-19 vaccines through routinely collected exposure and outcome using health registries’ [RS/2022/DTS/24104].

## Ethics

All study sites participating in this study conformed with their respective national and EU ethical and data protection requirements. Ethical statements for each of the participating study sites:

### Belgium

Data linkage and collection within the data-warehouse have been approved by the information security committee. The study was conducted in accordance with the Declaration of Helsinki. Ethical approval was granted for the gathering of data from hospitalised patients by the Committee for Medical Ethics from the Ghent University Hospital (reference number BC-07507) and authorisation for possible individual data linkage using the national register number from the Information Security Committee (ISC) Social Security and Health (reference number IVC/KSZG/20/384). Linkage of hospitalised patient data to vaccination and testing within the LINK-VACC project was approved by the Medical Ethics Committee UZ Brussels-VUB on 3 February 2021 (reference number 2020/523), and authorisation from the ISC Social Security and Health (reference number IVC/KSZG/21/034).

### Denmark

Only administrative register data was used for the study. According to Danish law, ethics approval is exempt for such research, and the Danish Data Protection Agency, which is dedicated ethics and legal oversight body, thus waives ethical approval for the study of administrative register data when no individual contact of participants is necessary, and only aggregate results are included as findings. The study is, therefore, fully compliant with all legal and ethical requirements, and there are no further processes available regarding such studies.

### Navarre (Spain)

The study was approved by Navarre’s Ethical Committee for Clinical Research, which waived the requirement of obtaining informed consent.

### Norway

Ethical approval was granted by Regional Committees for Medical and Health Research Ethics (REC) Southeast (reference number 122745). The Norwegian Institute of Public Health has performed a Data Protection Impact Assessment (DPIA) for Beredt C19.

### Portugal

The study received approval from the Ethical Committee and the Data Protection Officer of the Instituto Nacional de Saúde Doutor Ricardo Jorge. Given that data was irreversibly anonymised, the need for the participants’ informed consent was waived by the Ethical Committee.

### Italy

This study, based on routinely collected data, was not submitted for approval to an ethical committee because the dissemination of COVID-19 surveillance data was authorised by the Italian law N. 52 of 19 May 2022, following the law decree N. 24 of 24 March 2022 (Article n. 13). Based on the same acts, the information on COVID-19 vaccination was retrieved by the Italian National Institute of Health using data from the National Immunisation Information System of the Italian Ministry of Health. Because of the retrospective design and the large size of the population under study, in accordance with the Authorisation n. 9 released by the Italian data protection authority on 15 December 2016, the individual informed consent was not requested for the conduction of this study.

## Conflict of interest

Authors declare no competing interests.

## Author contributions

S Bacci, N Nicolay, J Humphreys, B Nunes, and S Monge conceived the study. J Humphreys, B Nunes, N Nicolay and S Monge conceived the methods. All authors from Public Health institutions at each study site were responsible for data management and analysis at the study site level. J Humphreys was responsible for pooling site level estimates. J Humphreys drafted the manuscript, with the help of B Nunes, A Nardone, and E Kissling. All authors contributed to the interpretation of the results and critically reviewed the manuscript. All authors approved the final version of this manuscript. All authors within the VEBIS-EHR working group made a substantial contribution to the conception or design of the work, critically revised the manuscript, provided their final approval of the version to be published, and agreed to be accountable for all aspects of the work.

## Notes

### Competing Interest Statement

The authors have declared no competing interest.

### Clinical Protocols

https://www.ecdc.europa.eu/en/publications-data/protocol-covid-19-vaccine-effectiveness-estimation-using-health-data-registries

### Author Declarations

Belgium: Data linkage and collection within the data-warehouse have been approved by the information security committee. The study was conducted in accordance with the Declaration of Helsinki. Ethical approval was granted for the gathering of data from hospitalised patients by the Committee for Medical Ethics from the Ghent University Hospital (reference number BC-07507) and authorisation for possible individual data linkage using the national register number from the Information Security Committee (ISC) Social Security and Health (reference number IVC/KSZG/20/384). Linkage of hospitalised patient data to vaccination and testing within the LINK-VACC project was approved by the Medical Ethics Committee UZ Brussels, VUB on 3 February 2021 (reference number 2020/523), and authorisation from the ISC Social Security and Health (reference number IVC/KSZG/21/034). Denmark: Only administrative register data was used for the study. According to Danish law, ethics approval is exempt for such research, and the Danish Data Protection Agency, which is dedicated ethics and legal oversight body, thus waives ethical approval for the study of administrative register data when no individual contact of participants is necessary, and only aggregate results are included as findings. The study is, therefore, fully compliant with all legal and ethical requirements, and there are no further processes available regarding such studies. Navarre (Spain): The study was approved by Navarres Ethical Committee for Clinical Research, which waived the requirement of obtaining informed consent. Norway: Ethical approval was granted by Regional Committees for Medical and Health Research Ethics (REC) Southeast (reference number 122745). The Norwegian Institute of Public Health has performed a Data Protection Impact Assessment (DPIA) for Beredt C19. Portugal: The study received approval from the Ethical Committee and the Data Protection Officer of the Instituto Nacional de Saude Doutor Ricardo Jorge. Given that data was irreversibly anonymised, the need for the participants informed consent was waived by the Ethical Committee. Italy: This study, based on routinely collected data, was not submitted for approval to an ethical committee because the dissemination of COVID-19 surveillance data was authorised by the Italian law N. 52 of 19 May 2022, following the law decree N. 24 of 24 March 2022 (Article n. 13). Based on the same acts, the information on COVID-19 vaccination was retrieved by the Italian National Institute of Health using data from the National Immunisation Information System of the Italian Ministry of Health. Because of the retrospective design and the large size of the population under study, in accordance with the Authorisation n. 9 released by the Italian data protection authority on 15 December 2016, the individual informed consent was not requested for the conduction of this study.

## References

1. Casey JA, Schwartz BS, Stewart WF, Adler NE. Using Electronic Health Records for Population Health Research: A Review of Methods and Applications. Annu Rev Public Health. 2016;37:61–81.

2. Tartof SY, Slezak JM, Fischer H, Hong V, Ackerson BK, Ranasinghe ON, et al. Effectiveness of mRNA BNT162b2 COVID-19 vaccine up to 6 months in a large integrated health system in the USA: a retrospective cohort study. The Lancet. 2021 Oct 16;398(10309):1407–16.

3. Barda N, Dagan N, Cohen C, Hernán MA, Lipsitch M, Kohane IS, et al. Effectiveness of a third dose of the BNT162b2 mRNA COVID-19 vaccine for preventing severe outcomes in Israel: an observational study. The Lancet. 2021 Dec;398(10316):2093–100.

4. Tang P, Hasan MR, Chemaitelly H, Yassine HM, Benslimane FM, Al Khatib HA, et al. BNT162b2 and mRNA-1273 COVID-19 vaccine effectiveness against the SARS-CoV-2 Delta variant in Qatar. Nat Med. 2021 Dec;27(12):2136–43.

5. Vasileiou E, Simpson CR, Shi T, Kerr S, Agrawal U, Akbari A, et al. Interim findings from first-dose mass COVID-19 vaccination roll-out and COVID-19 hospital admissions in Scotland: a national prospective cohort study. The Lancet. 2021 May 1;397(10285):1646–57.

6. Tessier E, Stowe J, Tsang C, Rai Y, Clarke E, Lakhani A, et al. Monitoring the COVID-19 immunisation programme through a National Immunisation Management System – England’s experience [Internet]. medRxiv; 2021 [cited 2025 Jan 7]. p. 2021.09.14.21263578. Available from: https://www.medrxiv.org/content/10.1101/2021.09.14.21263578v1

7. Nunes B, Rodrigues AP, Kislaya I, Cruz C, Peralta-Santos A, Lima J, et al. mRNA vaccine effectiveness against COVID-19-related hospitalisations and deaths in older adults: a cohort study based on data linkage of national health registries in Portugal, February to August 2021. Eurosurveillance. 2021 Sep 23;26(38):2100833.

8. Moustsen-Helms IR, Emborg HD, Nielsen J, Nielsen KF, Krause TG, Mølbak K, et al. Vaccine effectiveness after 1st and 2nd dose of the BNT162b2 mRNA Covid-19 Vaccine in long-term care facility residents and healthcare workers – a Danish cohort study [Internet]. medRxiv; 2021 [cited 2025 Jan 7]. p. 2021.03.08.21252200. Available from: https://www.medrxiv.org/content/10.1101/2021.03.08.21252200v1

9. Fabiani M, Ramigni M, Gobbetto V, Mateo-Urdiales A, Pezzotti P, Piovesan C. Effectiveness of the Comirnaty (BNT162b2, BioNTech/Pfizer) vaccine in preventing SARS-CoV-2 infection among healthcare workers, Treviso province, Veneto region, Italy, 27 December 2020 to 24 March 2021. Euro Surveill Bull Eur Sur Mal Transm Eur Commun Dis Bull. 2021;26(17).

10. Nordström P, Ballin M, Nordström A. Effectiveness of heterologous ChAdOx1 nCoV-19 and mRNA prime-boost vaccination against symptomatic Covid-19 infection in Sweden: A nationwide cohort study. Lancet Reg Health – Eur [Internet]. 2021 Dec 1 [cited 2025 Jan 7];11. Available from: https://www.thelancet.com/journals/lanepe/article/PIIS2666-77622100235-0/fulltext

11. Martínez-Baz I, Miqueleiz A, Casado I, Navascués A, Trobajo-Sanmartín C, Burgui C, et al. Effectiveness of COVID-19 vaccines in preventing SARS-CoV-2 infection and hospitalisation, Navarre, Spain, January to April 2021. Eurosurveillance. 2021 May 27;26(21):2100438.

12. Gier B de, Andeweg S, Joosten R, Schegget R ter, Smorenburg N, Kassteele J van de, et al. Vaccine effectiveness against SARS-CoV-2 transmission and infections among household and other close contacts of confirmed cases, the Netherlands, February to May 2021. Eurosurveillance. 2021 Aug 5;26(31):2100640.

13. Lanes S, Brown JS, Haynes K, Pollack MF, Walker AM. Identifying health outcomes in healthcare databases. Pharmacoepidemiol Drug Saf. 2015;24(10):1009–16.

14. Young JC, Conover MM, Funk MJ. Measurement error and misclassification in electronic medical records: methods to mitigate bias. Curr Epidemiol Rep. 2018 Dec;5(4):343–56.

15. Smedt TD, Merrall E, Macina D, Perez-Vilar S, Andrews N, Bollaerts K. Bias due to differential and non-differential disease-and exposure misclassification in studies of vaccine effectiveness. PLOS ONE. 2018 Jun 15;13(6):e0199180.

16. Bailie R, Bailie J, Chakraborty A, Swift K. Consistency of denominator data in electronic health records in Australian primary healthcare services: enhancing data quality. Aust J Prim Health. 2015;21(4):450–9.

17. Baum U, Kulathinal S, Auranen K. Exposure misclassification bias in the estimation of vaccine effectiveness. PLOS ONE. 2021 May 13;16(5):e0251622.

18. Sacco C, Manica M, Marziano V, Fabiani M, Mateo-Urdiales A, Guzzetta G, et al. The impact of underreported infections on vaccine effectiveness estimates derived from retrospective cohort studies. Int J Epidemiol. 2024 Apr 11;53(3):dyae077.

19. Groenwold RHH, Nelson DB, Nichol KL, Hoes AW, Hak E. Sensitivity analyses to estimate the potential impact of unmeasured confounding in causal research. Int J Epidemiol. 2010 Feb 1;39(1):107–17.

20. Remschmidt C, Wichmann O, Harder T. Frequency and impact of confounding by indication and healthy vaccinee bias in observational studies assessing influenza vaccine effectiveness: a systematic review. BMC Infect Dis. 2015 Oct 17;15(1):429.

21. Doll MK, Pettigrew SM, Ma J, Verma A. Effects of Confounding Bias in COVID-19 and Influenza Vaccine Effectiveness Test-Negative Designs Due to Correlated Influenza and COVID-19 Vaccination Behaviors [Internet]. medRxiv; 2021 [cited 2024 Dec 12]. p. 2021.10.22.21265390. Available from: https://www.medrxiv.org/content/10.1101/2021.10.22.21265390v1

22. Loiacono MM, Van Aalst R, Pokutnaya D, Mahmud SM, Nealon J. Methods to account for measured and unmeasured confounders in influenza relative vaccine effectiveness studies: A brief review of the literature. Influenza Other Respir Viruses. 2022;16(5):846–50.

23. Gianfrancesco MA, Goldstein ND. A narrative review on the validity of electronic health record-based research in epidemiology. BMC Med Res Methodol. 2021 Oct 27;21:234.

24. Farmer R, Mathur R, Bhaskaran K, Eastwood SV, Chaturvedi N, Smeeth L. Promises and pitfalls of electronic health record analysis. Diabetologia. 2018 Jun 1;61(6):1241–8.

25. Protocol for a COVID-19 vaccine effectiveness estimation using health data registries, VEBIS multi-country study - Version 2.0 [Internet]. 2024 [cited 2024 Dec 12]. Available from: https://www.ecdc.europa.eu/en/publications-data/protocol-covid-19-vaccine-effectiveness-estimation-using-health-data-registries

26. Monge S, Humphreys J, Nicolay N, Braeye T, Van Evercooren I, Holm Hansen C, et al. Effectiveness of XBB.1.5 Monovalent COVID-19 Vaccines During a Period of XBB.1.5 Dominance in EU/EEA Countries, October to November 2023: A VEBIS-EHR Network Study. Influenza Other Respir Viruses. 2024 Apr;18(4):e13292.

27. Kislaya I, Sentís A, Starrfelt J, Nunes B, Martínez-Baz I, Nielsen KF, et al. Monitoring COVID-19 vaccine effectiveness against COVID-19 hospitalisation and death using electronic health registries in ≥65 years old population in six European countries, October 2021 to November 2022. Influenza Other Respir Viruses. 2023 Nov;17(11):e13195.

28. Fontán-Vela M, Kissling E, Nicolay N, Braeye T, Van Evercooren I, Holm Hansen C, et al. Relative vaccine effectiveness against COVID-19 hospitalisation in persons aged ≥ 65 years: results from a VEBIS network, Europe, October 2021 to July 2023. Euro Surveill Bull Eur Sur Mal Transm Eur Commun Dis Bull. 2024 Jan;29(1):2300670.

29. Nunes B, Humphreys J, Nicolay N, Braeye T, Van Evercooren I, Holm Hansen C, et al. Monovalent XBB.1.5 COVID-19 vaccine effectiveness against hospitalisations and deaths during the Omicron BA.2.86/JN.1 period among older adults in seven European countries: A VEBIS-EHR network study. Expert Rev Vaccines. 2024 Dec 31;23(1):1085–90.

30. Soares P, Machado A, Nicolay N, Monge S, Sacco C, Hansen CH, et al. COVID-19 vaccine effectiveness in the paediatric population aged 5–17 years: a multicentre cohort study using electronic health registries in six European countries, 2021 to 2022. Eurosurveillance. 2025 Feb 27;30(8):2400450.

31. Chapter 10: Analysing data and undertaking meta-analyses [Internet]. [cited 2025 Mar 3]. Available from: https://training.cochrane.org/handbook/current/chapter-10

32. Lipsitch M, Tchetgen Tchetgen E, Cohen T. Negative Controls: A Tool for Detecting Confounding and Bias in Observational Studies. Epidemiology. 2010 May;21(3):383.

33. Hak E, Verheij TJM, Grobbee DE, Nichol KL, Hoes AW. Confounding by indication in non-experimental evaluation of vaccine effectiveness: the example of prevention of influenza complications. J Epidemiol Community Health. 2002 Dec;56(12):951–5.

34. Jackson LA, Jackson ML, Nelson JC, Neuzil KM, Weiss NS. Evidence of bias in estimates of influenza vaccine effectiveness in seniors. Int J Epidemiol. 2006 Apr;35(2):337–44.

35. Simonsen L, Taylor RJ, Viboud C, Miller MA, Jackson LA. Mortality benefits of influenza vaccination in elderly people: an ongoing controversy. Lancet Infect Dis. 2007 Oct 1;7(10):658–66.

36. Jackson ML, Yu O, Nelson JC, Naleway A, Belongia EA, Baxter R, et al. Further Evidence for Bias in Observational Studies of Influenza Vaccine Effectiveness: The 2009 Influenza A(H1N1) Pandemic. Am J Epidemiol. 2013 Oct 15;178(8):1327–36.

37. Castilla J, Guevara M, Martínez-Baz I, Ezpeleta C, Delfrade J, Irisarri F, et al. Enhanced Estimates of the Influenza Vaccination Effect in Preventing Mortality: A Prospective Cohort Study. Medicine (Baltimore). 2015 Jul;94(30):e1240.

38. Chemaitelly H, Ayoub HH, Coyle P, Tang P, Hasan MR, Yassine HM, et al. Assessing Healthy Vaccinee Effect in COVID-19 Vaccine Effectiveness Studies: A National Cohort Study in Qatar [Internet]. medRxiv; 2024 [cited 2025 Jan 8]. p. 2024.07.28.24311115. Available from: https://www.medrxiv.org/content/10.1101/2024.07.28.24311115v2

39. Hansen CH, Moustsen-Helms IR, Rasmussen M, Søborg B, Ullum H, Valentiner-Branth P. Short-term effectiveness of the XBB.1.5 updated COVID-19 vaccine against hospitalisation in Denmark: a national cohort study. Lancet Infect Dis. 2024 Feb 1;24(2):e73–4.

40. Poukka E, Auranen K, Baum U. Non-COVID-19 hospitalisation as a negative control outcome in COVID-19 vaccine effectiveness studies. Lancet Infect Dis. 2024 May 1;24(5):e275.

41. Hansen CH, Moustsen-Helms IR, Rasmussen M, Søborg B, Ullum H, Valentiner-Branth P. Non-COVID-19 hospitalisation as a negative control outcome in COVID-19 vaccine effectiveness studies – Authors’ reply. Lancet Infect Dis. 2024 May 1;24(5):e276.

42. European Health Data Space - European Commission [Internet]. 2024 [cited 2024 Oct 31]. Available from: https://health.ec.europa.eu/ehealth-digital-health-and-care/european-health-data-space_en

43. Framework for financial data access - European Commission [Internet]. [cited 2025 Mar 5]. Available from: https://finance.ec.europa.eu/digital-finance/framework-financial-data-access_en

44. Double Negative Control Inference in Test-Negative Design Studies of Vaccine Effectiveness - PMC [Internet]. [cited 2025 Apr 30]. Available from: https://pmc.ncbi.nlm.nih.gov/articles/PMC8963685/

45. Martens EP, Pestman WR, de Boer A, Belitser SV, Klungel OH. Instrumental variables: application and limitations. Epidemiol Camb Mass. 2006 May;17(3):260–7.

46. Brookhart MA, Rassen JA, Schneeweiss S. Instrumental variable methods in comparative safety and effectiveness research. Pharmacoepidemiol Drug Saf. 2010 Jun;19(6):537–54.

47. Bang H, Robins JM. Doubly robust estimation in missing data and causal inference models. Biometrics. 2005 Dec;61(4):962–73.

48. Funk MJ, Westreich D, Wiesen C, Stürmer T, Brookhart MA, Davidian M. Doubly Robust Estimation of Causal Effects. Am J Epidemiol. 2011 Apr 1;173(7):761–7.

49. Self controlled case series methods: an alternative to standard epidemiological study designs | The BMJ [Internet]. [cited 2025 Mar 5]. Available from: https://www.bmj.com/content/354/bmj.i4515

